# Assessment of Fairness and Bias of an Image-based Surgical Site Infection Detection AI Model

**DOI:** 10.64898/2026.09.22.26363636

**Authors:** Frank G. Lee, Ashok Choudhary, Leo Y. Li-Han, Erica L. Dale, Audrey C. Bankes, Chris Varghese, Stephanie S. Anderson, Elizabeth B. Habermann, David A. Etzioni, Sarah A. McLaughlin, Hojjat Salehinejad, Cornelius A. Thiels

## Abstract

**Background:** Evaluating fairness in artificial intelligence (AI) is essential before clinical implementation to prevent unintended harm across diverse patient populations. This study assesses performance differences of an image-based surgical site infection (SSI) detection model across age, gender, geography, race, and skin color.

**Methods:** Model development/validation study using retrospective, multicenter data across surgical subspecialties from adults who submitted an image of their incision via the electronic health record within 30 days of their procedure between 2019-2024. Area under receiver operating curve (AUROC) was compared by age (<55 vs. ≥55 years old), gender (male vs. female), geography (Rochester, Minnesota vs. Jacksonville, Florida vs. Scottsdale, Arizona vs. regional Minnesota/Wisconsin areas), race (White/Caucasian vs. non-White/Caucasian), and skin color (Fitpatrick I-III vs. IV-VI and Monk A-E vs. F-J).

**Results:** Training data contained 13,702 images from 4,274 patients between 2019-2022 (50.1% ≥55 years old; 63.2% female; 91.6% White/Caucasian; 58.5% Rochester, 17.3% Florida, 10.5% Arizona; 96.3% light skin tone), of which 1,348 images (9.8%) had an SSI. No differences in SSI detection existed for age, gender, race, or skin color (AUROC 0.83-0.87, p>0.09), however geographical differences were significant: lowest AUROC 0.83 (95% CI 0.77-0.88) vs. highest AUROC 0.87 (95% CI 0.84-0.89, p=0.01). Temporal validation using 4,242 images from 1,353 patients between 2022-2023 showed intact fairness across subgroups (ΔAUROC ≤0.06) except geography (ΔAUROC ≤0.16).

**Conclusions:** The model behaved fairly across age, gender, race, and skin color, however geographical differences persisted. These findings have practical implications underscoring the importance of evaluating bias in clinical AI before deployment.

## Introduction

Bias and fairness evaluation of artificial intelligence (AI) technologies are essential before clinical implementation to ensure equitable performance across diverse patient populations and to prevent unintended harm. The risk of deploying biased AI is evident in real-world examples of policing,^1^ criminal sentencing,^2^ and healthcare.^3^ If left unexplored and unmitigated, bias can negatively impact fairness resulting in systematic disadvantage against particular groups especially those with protected attributes: characteristics which have historically been a source of discrimination and currently have legal safeguards in place e.g. age, sex/gender, race/ethnicity, socioeconomic status, and nationality.^4^ This is especially important when sociodemographic attributes and proxies including geography and skin color can perpetuate inequity and erode patient trust.^5^ Systematic reviews have shown bias, fairness, and equity analyses are seldom performed in <12% AI model development/evaluation studies, and when performed, are often limited to a single sociodemographic factor usually race.^6–8^

AI has significant potential in assessment of wounds with over 330 million surgeries performed annually.^9,10^ Patients increasingly send images to their healthcare teams via mobile digital health platforms to surveil postoperative incisions. Delays in care can have significant consequences particularly for surgical site infections (SSIs) which remains the costliest healthcare-associated infection.^11^ To address this problem, we developed an AI-enabled postoperative wound assessment tool which analyzes patient-submitted images to improve triage and detection of SSIs.^12^ However, the fairness of these models remains unknown, particularly on skin color.^13–15^ Therefore, we comprehensively evaluated the fairness, as a measure of accuracy and consistent performance across subgroups, of our computer vision SSI detection model for key sociodemographic factors and skin color. Herein, we provide a practical case study for evaluating equity-based fairness for an image-based surgical AI tool.

## Methods

### Study Design

This was a model evaluation study using retrospective data. This study was approved by the Mayo Clinic Institutional Review Board (#23-013072) and reported in accordance with TRIPOD+AI.^16^ All procedures were performed in compliance with relevant laws and institutional guidelines without patient or public involvement.

### Data Source and Image Acquisition

The training cohort consisted of adult patients (≥18 years old) who had surgery at one of the 9 Mayo Clinic hospitals across Arizona, Florida, Minnesota, and Wisconsin between January 1, 2019 and December 31, 2022. Inclusion criteria were patients who had submitted an image (file format: .jpg, .jpeg, .png) of their postoperative wound/incisions to the electronic health record via the patient portal within 30 days of their procedure. Only surgical cases with abstracted National Surgical Quality Improvement Program (NSQIP) outcomes data were included. Procedures from all surgical subspecialties were included except for trauma, cardiac, and transplant which are not captured in NSQIP. All images with associated date/time stamp and patient information were downloaded using Structured Query Language (SQL).

### Variables and Definitions

Collected variables included age at surgery, gender (as listed in the electronic medical record), self-reported race, procedure, date of surgery, surgical subspecialty, hospital, and NSQIP outcomes for superficial and deep SSI. NSQIP collects outcomes up to 30 days after surgery and defines SSI based on Center for Disease and Control (CDC) guidelines.^17^ Hospitals were grouped by geographical region. These groupings were: Rochester, Minnesota (RST); Scottsdale, Arizona (ARZ); Jacksonville, Florida (FLA); and Mayo Clinic Health System (MCHS) comprising regional/rural Minnesota and Wisconsin. Both a positive NSQIP diagnosis for superficial and/or deep SSI *and* visible evidence of SSI in the image per clinician review served as the ground truth for SSI outcome during model development. Organ-space SSI was not used as these infections are deep and not visible to the naked eye.

### Image Review

Only images with incisions were used for bias and fairness evaluation, as images without incisions were not applicable for sociodemographic and skin color assessment. There was no minimum resolution requirement. All images were initially screened by a surgeon (F.L.) for visible incision irrespective of closure type e.g. sutures, staples, or secondary intention. Images without incisions included paper forms, medical equipment, dressings, and skin rashes (n=10,468). Images containing incisions from patients who had a positive diagnosis for superficial/deep SSI in NSQIP were manually reviewed by two surgeons (F.L. and H.M.) for visible evidence of SSI within the image and classified into binary categories of presence (yes vs. no). Inter-rater concordance had substantial agreement (Cohen’s kappa 0.63).^18^ Disagreements were resolved by discussion, and a third senior surgeon (C.T.) was involved as necessary for ties.

Skin color ratings were based on Fitzpatrick and Monk scales. Skin color was reviewed by two dermatology trainees (E.D. and A.B.) in duplicate for images with skin incision present. An estimated Fitzpatrick rating was assigned based on clinician assessment of skin available in the image, which has been previously described.^13,19^ Fitzpatrick scale consists of 6 skin tone levels (I-VI)^20^; Monk contains 10 levels (A-J).^21^ Each scale was grouped into light (Fitzpatrick I-III or Monk A-E) and dark skin color (Fitzpatrick IV-VI or Monk F-J). A reference guide for skin color rating assignment is available in **Figure A.1**. Inter-rater concordance for Fitzpatrick and Monk light and dark groupings had substantial agreement (Cohen’s kappa 0.70 and 0.68 respectively).^18^ Disagreements were resolved by a third clinician (F.L.).

### Model Development

Training of the deep learning model has been previously described by the original study detailing model development.^12^ A Vision Transformer architecture was used. As an overview, Vision Transformer (vit_large_patch16_224) converts an image into a grid of 196 smaller patches (grid of 14×14), and the patches are linearized into a single array with spatial relationship information encoded by a position embedding layer (**Methods A.2**).^22^ The model was trained to classify images with visible SSI versus images with normal-appearing incision. Due to variations in image height-width size ratio, a pre-processing step added padding to standardize all image dimensions to 224×224 pixels. Preliminary experiments were performed to determine ideal configuration settings for hyperparameters such as batch size (the number of images used to update the model parameters per iteration) and learning rate (the rate which the model is updated and adjusts weights during training) (**Table A.3**). A batch size of 32 with a learning rate of 1×10^-5^ provided the highest validation accuracy. For model development, 5-fold cross-validation was used (i.e. 80/20 training/validation split where 80% of applicable images were used for training and remaining 20% were reserved for internal validation testing, then repeated 4 times for a total of 5 folds, each with different partitions of training images). For training, the number of SSI examples was randomly upsampled 1:1 to balance the non-SSI images to address class imbalance. Explainable AI (XAI) experiments were conducted using gradient-weighted class activation mapping (GradCAM) to create attention heatmaps to ensure the image area of the wound was used for infection prediction versus proxy artifacts e.g. demarcations by skin marker, rulers, or background content (**Figure 1**). Model training occurred between September 2025 and April 2026.

**Figure 1:**
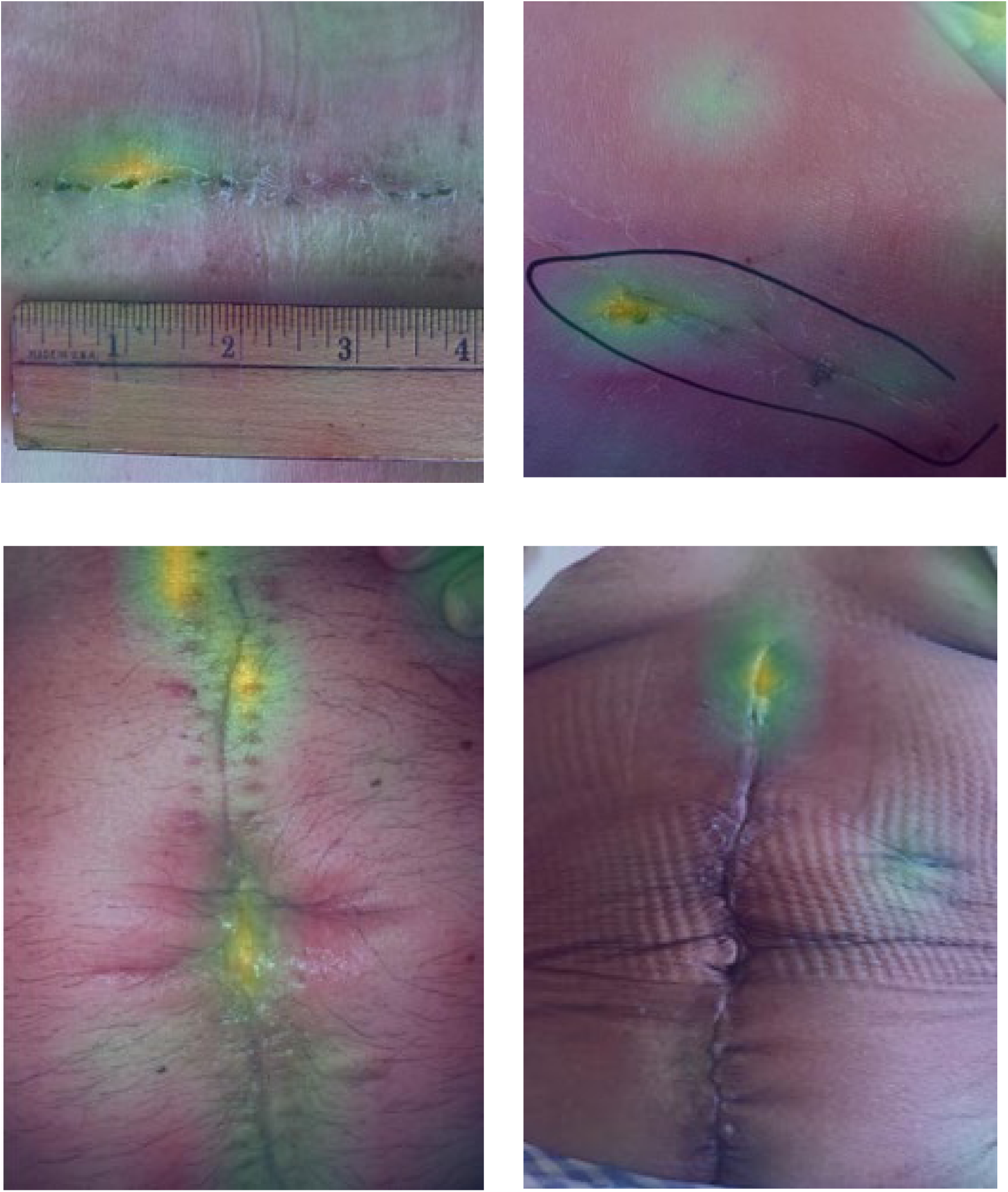
Example patient-submitted images overlaid with model attention heatmaps (lighter green areas weighted higher importance for model prediction)

### Model Input, Output, and Intended Use

The only predictor input to the model was the image, and the model was trained to the output of SSI diagnosis per NSQIP and human reviewers. For each image input, the model generated an estimated probability for the outcome (i.e. SSI) on a continuous scale between 0 (no risk) to 1 (highest likelihood). The cut-off threshold for determining positive prediction based on the continuous scale was analytically derived (see Data Analysis). The intended use of this model is for screening/triaging purposes and not intended to replace clinician judgment or be diagnostic. The issue of low-image quality (e.g. blurriness, low lighting, obstructed view, zoomed in/out) was discussed in the original development paper, and a quality check was implemented to ensure minimum acceptable image quality to enable appropriate evaluation for incision and infection.^12^ These challenges limited characterization of the incision e.g. length or surface area.

### Bias and Fairness Subgroup Analysis

Subgroups of interest for bias and fairness evaluation were based on age, gender, geography, race, and skin color. These subgroups were defined as follows: age <55 and ≥55; female and male; RST, FLA, ARZ, and MCHS campuses; White/Caucasian and non-White/Caucasian; and light and dark skin color using Fitzpatrick and Monk scales. Non-binary gender [n=10 images (0.07%) from 3 patients] and unknown/not reported race [n=412 images (3.0%) from 114 patients] were omitted. Equity analysis was performed during cross-validation with the validation test set comprising only images from the subgroup of interest. For validation, a natural distribution (no balancing methods applied) was used as this reflects real-world settings and produces the most representative results.^23–25^

### Temporal Validation

Temporal validation was performed using images from January 1, 2023 to January 31, 2024. All images were submitted within 30 days of surgery with cases associated with NSQIP SSI outcomes. Images were manually reviewed for visible incision, signs of infection, and assigned skin color ratings using the same procedures above and reviewers (F.L., A.B., E.D.). Images were categorized into the same subgroupings for age, gender, geography, race, and skin color. The model was run on images containing incisions for detecting SSI within each subgroup arm using a natural distribution without correction of class imbalance. Temporal validation testing occurred between December 2025 and April 2026.

### Statistical Analysis

Model performance was evaluated using area under receiver operating curve (AUROC) for each subgroup as mean ± standard deviation (SD) across all folds. Bootstrapped t-test compared subgroup AUROC within a class. For geography (with four subgroups), a one-vs-all approach was used. Significance was defined as p<0.05. All statistical tests were two-sided. Additional performance metrics included accuracy, precision (positive predictive value, PPV), recall (sensitivity), specificity, and F1-score (see **Equations A.4**). For threshold-dependent metrics, the threshold which maximized F1-score (ideal for imbalanced datasets) was used and reported in the results. For binary classification fairness within subgroups, predictive parity (equal PPV) and equalized odds (equal true positive rate and equal false positive rate) were assessed. Descriptive statistics summarized sociodemographic variables using median and interquartile range (IQR). Sample size determination was based on inclusion of all available training data. Temporal cut-offs for development/evaluation datasets were informed by available NSQIP outcomes data at time of model development and evaluation. Missing values were omitted if missingness comprised <5% of the variable of interest, otherwise missing values were treated as their own category. No imputations were performed for missingness. Statistical software included PyTorch version 2.10. Statistical Analysis System (SAS) version 9.4 (SAS Institute Inc., Cary, North Carolina) was used for querying wound image attachments.

## Results

The training dataset included 4,274 postoperative patients. Median age was 54 (IQR 41-65); 2,630 (61.5%) females; 3,943 (92.3%) White/Caucasian, 108 (2.5%) Black/African American, 72 (1.7%) Asian, 37 (0.9%) Alaska Native/Native Hawaiian/Other, and 114 (2.7%) unknown/not reported. Patients were predominantly treated in Rochester, Minnesota (2,222, 52.0%), followed by Florida (715, 16.7%) and Arizona (534, 12.5%). Each patient submitted a median of 2 (IQR 1-4) images for a total of 13,702 photos with a visible incision, and 1,348 (9.8%) containing SSI. NSQIP SSI classification type were 1,244 (92.3%) superficial, 96 (7.1%) deep, and 8 (0.6%) both. Skin color labelling of images revealed predominantly light skin representation (94.7% Fitzpatrick I-III and 96.3% Monk A-E) and few dark skin examples (5.3% Fitzpatrick IV-VI and 3.7% Monk F-J). Complete sociodemographic and skin color characteristics are available in **Table 1**. Surgical case mix across geographical campuses varied with greater proportion of orthopedic surgery in Florida (37.4%), plastic surgery in Rochester (29.7%), thoracic surgery in Arizona (10.7%), and obstetrics in the health systems (9.8%) (**Table A.5**).

**Table 1:** Overview of characteristics in training dataset.

|  | Photos containing incision | Photos containing SSI |
| --- | --- | --- |
| Count | 13,702 (100%) | 1,348 (9.8%) |
| **Age** |  |  |
| <55 years old | 6,837 (49.9%) | 645 (9.4%) |
| ≥55 years old | 6,865 (50.1%) | 703 (10.2%) |
| **Gender** |  |  |
| Female | 8,659 (63.2%) | 852 (9.8%) |
| Male | 5,033 (36.7%) | 496 (9.9%) |
| **Race** |  |  |
| White/Caucasian | 12,545 (91.6%) | 1276 (10.2%) |
| Black/African American | 367 (2.7%) | 34 (9.3%) |
| Asian | 259 (1.9%) | 18 (6.9%) |
| Other | 119 (0.9%) | 3 (2.5%) |
| Unknown | 412 (3.0%) | 17 (4.1%) |
| **Skin Color** |  |  |
| Fitzpatrick |  |  |
| Light | 12,973 (94.7%) | 1,287 (9.9%) |
| Dark | 729 (5.3%) | 61 (8.4%) |
| Monk |  |  |
| Light | 13,190 (96.3%) | 1,299 (9.8%) |
| Dark | 512 (3.7%) | 49 (9.6%) |
| **Geography** |  |  |
| Rochester | 8,009 (58.5%) | 789 (9.9%) |
| Florida | 2,377 (17.3%) | 197 (8.3%) |
| Minnesota/Wisconsin | 1,872 (13.7%) | 214 (11.4%) |
| Arizona | 1,444 (10.5%) | 148 (10.2%) |

Values provided as N (%). Row percentages are provided for photos containing SSI column. Other race includes Alaska Native, Hawaiian Native, and multiple race. SSI surgical site infection.

### Bias and Fairness Subgroup Analysis

Bias and fairness evaluation of the SSI detection model showed no differential performance across age, gender, race, or skin color during model development (**Figure 2**). Overall model performance across folds had a mean ± SD AUROC of 0.854 ± 0.004. The model exhibited a statistically significant difference in performance for geography, with the lowest detection performance for patients treated in Florida versus highest performance for those treated in Rochester, Minnesota (mean AUROC 0.828 vs. 0.865, p=0.010). However, all subgroups achieved a minimum performance of 0.8 and subgroup-specific differences were <0.04, suggesting acceptable level of fairness across geographical subgroups. For race, non-White/Caucasian had equivalent performance to White/Caucasian (0.865 ± 0.035 vs. 0.852 ± 0.003, p=0.582) despite class imbalance (55/774 vs. 1266/12477 photos with SSIs). Similarly, dark skin color had equivalent performance compared to light skin color for Fitzpatrick (0.874 ± 0.043 vs. 0.852 ± 0.004, p=0.251) and Monk (0.886 ± 0.014 vs. 0.853 ± 0.004, p=0.292) skin color scales despite class imbalance (with 61/728 Fitzpatrick and 49/511 Monk positive SSI examples within the dark skin subgroup) (**Table 2**). Non-White/Caucasian and dark skin tone patients had higher PPV than White/Caucasian and light skin tone patients (66-73% vs. 50%), however all subgroups achieved a PPV of 50% except Florida geography (42.4%), suggesting a lack of predictive parity in this subgroup. Age, gender, and race had equivalent true positive rates and false positive rates, supporting the notion of equalized odds (**Table 3**). Lower true positive rates were observed in patients with light skin color and those from Florida and Arizona geographical regions (confusion matrices available in **Figure A.6**). Temporal validation on images submitted between 2023-2024 [n=4,242 images from 1,353 patients; median age 55 (IQR 41-66); 863 (63.8%) female; 1,235 (91.3%) White/Caucasian] demonstrated overall AUROC 0.680. Characteristics between the training and validation datasets were similar except a lower rate of SSI among images for the newer cohort: 9.8% (2019-2022) vs. 6.5% (2023-2024) (**Table 4**). Fairness on temporal validation showed AUROC subgroup differences ranging from 0.01 for Fitzpatrick skin color class to 0.16 for geography class (**Figure 3**). Except geography, all subgroups were within AUROC +/-0.06 difference supporting intact fairness on temporal validation. Additional details on temporal validation fairness metrics are provided in **Table A.7.**

**Figure 2:**
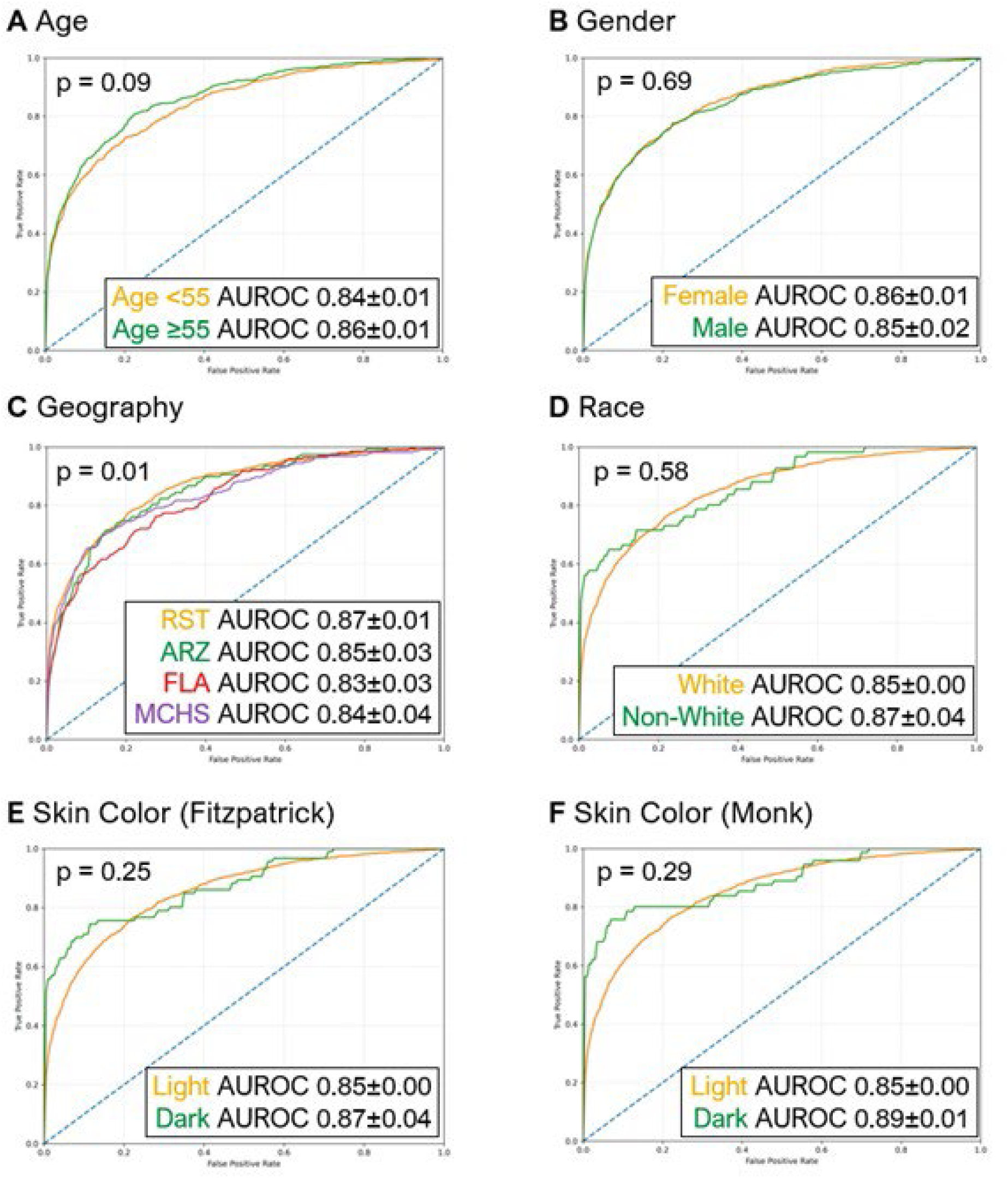
Development subgroup comparison of SSI detection AUROC model performance by (A) age, (B) gender, (C) geography, (D) race, (E) skin color by Fitzpatrick scale, and (F) skin color by Monk scale. Values reported as mean ± SD.

**Figure 3:**
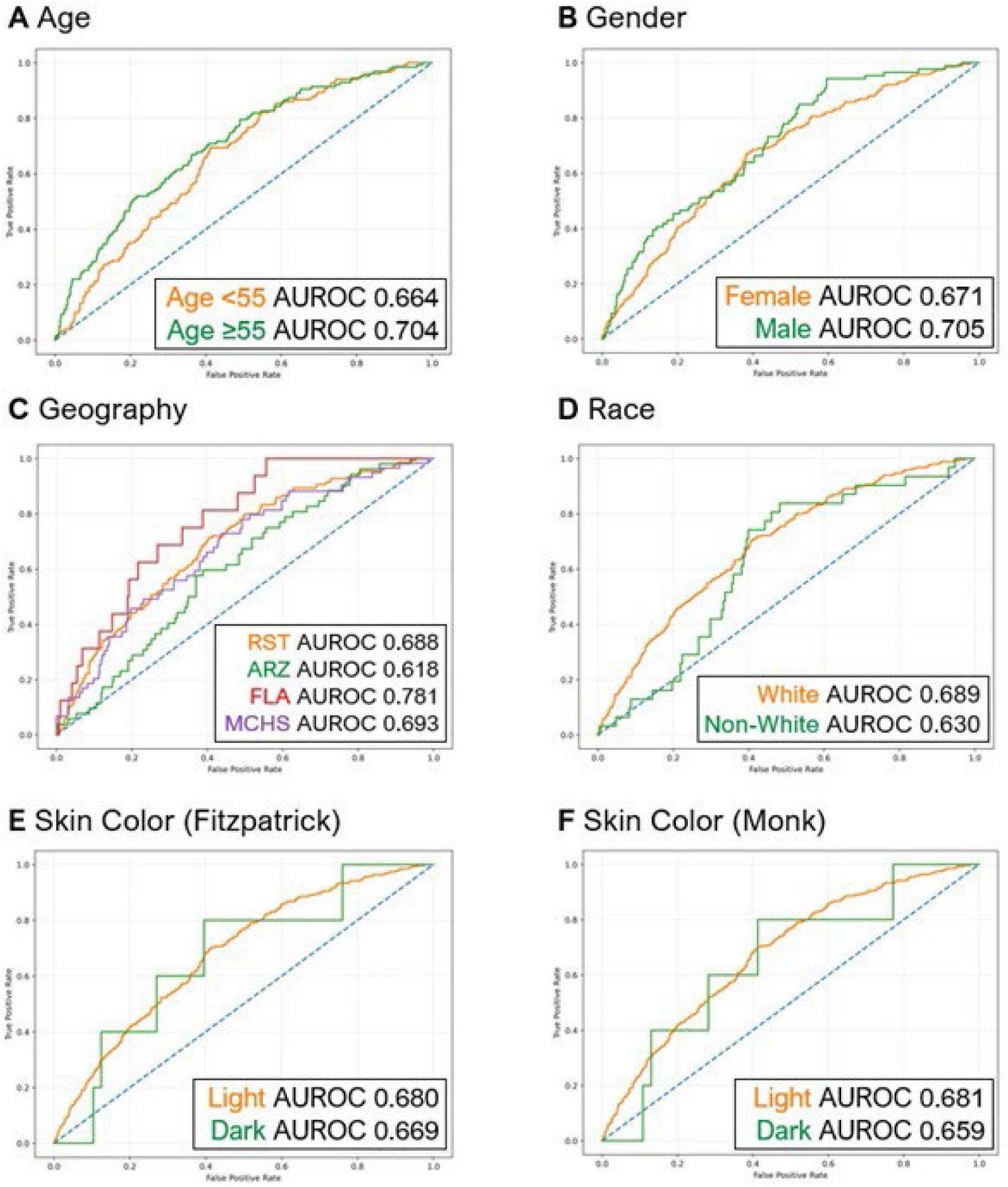
Temporal validation of SSI detection for AUROC by subgroups of (A) age, (B) gender, (C) geography, (D) race, (E) skin color by Fitzpatrick scale, and (F) skin color by Monk scale. Values reported as mean ± SD.

**Table 2:**
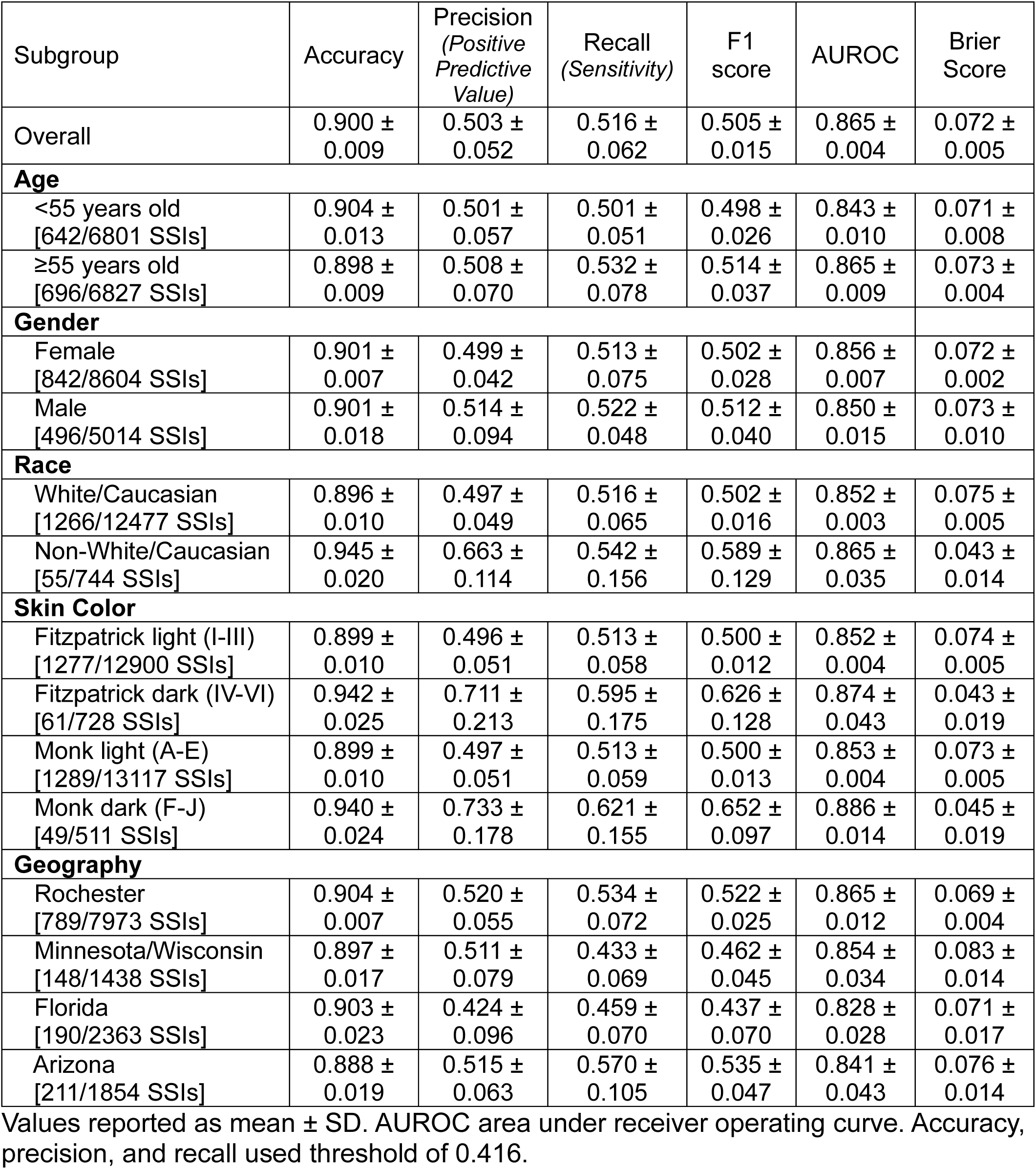
SSI detection performance metrics by subgroup during model development.

**Table 3:** True positive and true negative rates for subgroup equalized odds analysis during model development.

|  | TP | FP | FN | TN | TPR | FPR |
| --- | --- | --- | --- | --- | --- | --- |
| **Age** |  |  |  |  |  |  |
| <55 years old | 323 | 333 | 319 | 5826 | 50.3% | 94.6% |
| ≥55 years old | 368 | 370 | 328 | 5761 | 52.9% | 94.0% |
| **Gender** |  |  |  |  |  |  |
| Male | 431 | 442 | 411 | 7320 | 51.2% | 94.3% |
| Female | 260 | 261 | 236 | 4257 | 52.4% | 94.3% |
| **Race** |  |  |  |  |  |  |
| White/Caucasian | 653 | 680 | 613 | 10531 | 51.6% | 93.9% |
| Non-White/Caucasian | 29 | 16 | 26 | 673 | 52.7% | 97.7% |
| **Skin Color** |  |  |  |  |  |  |
| Fitzpatrick light (I-III) | 655 | 686 | 622 | 10937 | 51.3% | 94.1% |
| Fitzpatrick dark (IV-VI) | 36 | 17 | 25 | 650 | 59.0% | 97.5% |
| Monk light (A-E) | 661 | 691 | 628 | 11137 | 51.3% | 94.2% |
| Monk dark (F-J) | 30 | 12 | 19 | 450 | 61.2% | 97.4% |
| **Geography** |  |  |  |  |  |  |
| Rochester | 421 | 400 | 368 | 6784 | 53.4% | 94.4% |
| Minnesota/Wisconsin | 120 | 118 | 91 | 1525 | 56.9% | 92.8% |
| Florida | 87 | 123 | 103 | 2050 | 45.8% | 94.3% |
| Arizona | 63 | 62 | 85 | 1228 | 42.6% | 95.2% |

Reported using threshold of 0.416. TP true positive, FP false positive, FN false negative, TN true negative, TPR true positive rate, FPR false positive rate.

**Table 4:** Overview of characteristics in temporal validation dataset.

|  | Photos containing incision | Photos containing SSI |
| --- | --- | --- |
| Count | 4,242 (100%) | 277 (6.5%) |
| **Age** |  |  |
| <55 years old | 2,056 (48.5%) | 150 (7.3%) |
| ≥55 years old | 2,186 (51.5%) | 127 (5.8%) |
| **Gender** |  |  |
| Female | 2,821 (66.5%) | 191 (6.8%) |
| Male | 1,421 (33.5%) | 86 (6.1%) |
| **Race** |  |  |
| White/Caucasian | 3,784 (89.2%) | 245 (6.5%) |
| Black/African American | 191 (4.5%) | 5 (2.6%) |
| Asian | 128 (3.0%) | 1 (0.8%) |
| Other | 85 (2.0%) | 25 (29.4%) |
| Unknown | 54 (1.3%) | 1 (1.9%) |
| **Skin Color** |  |  |
| Fitzpatrick |  |  |
| Light | 4,067 (95.9%) | 272 (6.7%) |
| Dark | 175 (4.1%) | 5 (2.9%) |
| Monk |  |  |
| Light | 4,073 (96.0%) | 272 (6.7%) |
| Dark | 169 (4.0%) | 5 (3.0%) |
| **Geography** |  |  |
| Rochester | 2,259 (53.3%) | 150 (6.6%) |
| Florida | 791 (18.7%) | 59 (7.5%) |
| Minnesota/Wisconsin | 673 (15.9%) | 52 (7.7%) |
| Arizona | 519 (12.2%) | 16 (3.1%) |

Values provided as N (%). Row percentages are provided for photos containing SSI column. Other race includes Alaska Native, Hawaiian Native, and multiple race. SSI surgical site infection.

## Discussion

An AI postoperative wound assessment tool was evaluated for bias which demonstrated equitable performance for skin color and protected sociodemographic attributes like age, gender, and race. Interestingly, geographical variation demonstrated statistically significant differences in AUROC performance which may have important implications before deployment, potentially requiring recalibration or retraining. Possible explanations for the geographical differences may be practice-specific patterns in how patients are instructed to send in images or compositional variation in surgical case mix. Overall, the impact of geographical difference was minimal. All geographical subgroups had acceptable minimum metrics limiting the notion of systematic disadvantage or inequity. The temporal validation showed a reduction in performance, which is to be expected with a completely naïve and different dataset, but importantly fairness was intact for all subgroups except geography. Future external and prospective validation studies are needed to establish generalizability, clinical effectiveness, and impact on patient outcomes.

Examples of implicit learning can have unintended consequences on model performance.^26,27^ Dehkharghanian et al. demonstrated how a pathomics model trained to diagnose the type of cancer from de-identified histology slides was able to identify the originating institution – through slide artifacts and staining techniques potentially introducing a shortcut bias based on site-specific patterns rather than true pathology.^26^ Our findings showing geographical differences in SSI detection further underscore the importance of geography (or acquisition site or location) as a subgroup class for evaluating fairness. There are adjuncts to combat implicit learning during development like GradCAM attention heatmaps which can be a useful check for unintended features.

As exploration of predictive AI applications grows, efforts to measure and mitigate algorithmic bias should be performed but optimal strategy is evolving: predictive AI development has become facile and rapid, while evaluation and validation has necessitated careful review. Systematic reviews and meta-analyses show that bias, fairness, and equity analyses are being underperformed.^6–8^ Reporting guidelines have been updated with heavy emphasis on AI fairness achieving as fair as possible performance across groups, including but not limited to “sex, gender, age, ethnicity, socioeconomic status, medical conditions, and acquisition site [geography].”^16,23,28^ Selection of appropriate measures is necessary in fairness analysis: AUROC is preferred over AUPRC,^4^ and predictive parity and equalized odds are recommended in binary classification tasks to ensure minority groups or those with protected attributes are not missed or given unequal opportunities.^29,30^ The fairness of binary classification will be highly dependent upon threshold selection.

For AI models involving images of skin, there is a lack of consensus on which skin color scale should be used.^31,32^ Both dermatology and AI literature increasingly recommend discontinuing the use of Fitzpatrick due to poor diversity and reliability.^33,34^ Yet Fitzpatrick scale remains widely used: a systematic review of deep learning algorithms for skin disease showed Fitzpatrick is the main reporting measure for skin color assessment.^14^ As originally devised, the Fitzpatrick scale combined clinician assessment with patient-reported frequency of sunburning.^20^ Recently, modified Fitzpatrick labeling has been performed without photosensitivity information by relying on consensus from multiple clinicians, as conducted in this present study, however these modifications may sacrifice validity of the original Fitzpatrick measure.^13,19^ Rochon et al. showed decreased sensitivity in Fitzpatrick dark skin tones for detecting skin separation and redness in postoperative wound images. Their model used a You Only Look Once (YOLO) architecture, ideal for mobile and edge devices because of its lower computational requirement, as opposed to Vision Transformer which is state-of-the-art, computationally intensive, and has greater global spatial awareness and self-attention.^9,35^

The low number of SSI examples in all patients, but especially minority groups like non-White/Caucasian patients and patients with dark skin tones, is a limitation and presents a barrier for a fully robust fairness evaluation. Other recently developed AI-based wound image SSI detection models lack demographic information or were validated on only White patients.^9,36^ The STANDING Together Initiative aims for diverse and balanced representation in datasets.^37^ We support this initiative and further monitoring of our model is required as more diverse datasets become increasingly available to mitigate a type II error. The lack of representation in our dataset is not unique; the Gender Shades study showed public datasets suffer from a similar imbalance: in their analysis, 77.5% were male and 83.5% White/Caucasian.^38^ A resulting challenge is class imbalance between the majority and minority subgroups. We utilized a balanced training dataset and imbalanced (i.e. natural distribution) validation dataset to pragmatically reflect real-world settings, following best practice recommendations which found class imbalance correction can lead to miscalibration and overestimation of true risk.^23–25^ The decision to balance or not balance is context- and scenario-dependent with potential trade-offs between performance gain and fairness loss.^39^

A strength of this study is the large number of clinical photos not from a publicly available benchmark dataset. Within our institution, approximately 140,000 major surgeries are performed each year with patients submitting 35,000 postoperative images annually. The administrative burden of reviewing these images promptly and risk of treatment delays is high, and an opportunity for a more intelligent path forward may exist with our proposed model. Our model attempts to address other forms of bias and disparities, including cognitive (variability in provider assessment) and access (patients who live in medical deserts or have limited transportation to be physically seen by a provider). As computer vision foundation models and camera image quality continue to evolve, the opportunities ahead are promising with greater advancements in performance and accuracy. Continued iteration and model maturation is necessary.

In conclusion, fairness of an image-based AI model trained to detect infections in the setting of postoperative wound monitoring was demonstrated for age, gender, race, and skin color. Despite geographical differences, all subgroups achieved a minimum acceptable performance threshold. This study is an important step in evaluating clinical AI tools before clinical deployment and serves as a practical case study for assessing sociodemographic and skin color bias in a surgical wound monitoring tool.

## Data Availability

Due to nature of images and patient privacy as it relates to HIPAA, study data cannot be shared.

## Acknowledgments

Special thanks to Dr. Hala Muaddi for her assistance with reviewing wound images during the initial phase of this work.

## Data Sharing Statement

Due to nature of images and patient privacy as it relates to HIPAA, study data cannot be shared.

## Code Availability

The underlying code for this study and training/validation datasets are not publicly available but may be made available to qualified researchers on reasonable request to the corresponding author.

## Authors’ Contributions

Frank Lee wrote the initial draft of the manuscript. Ashok Choudhary and Leo Li-Han wrote the code and ran the experiments to train/test the model. Frank Lee, Erica Dale, and Audrey Bankes labelled photos. Chris Varghese assisted with study design. Stephanie Anderson acquired and compiled all images for research. Elizabeth Habermann, David Etzioni, and Sarah McLaughlin approved staff and image resourcing. Cornelius Thiels acquired grant funding. Hojjat Salehinejad and Cornelius Thiels supervised and were responsible for conception of the study. All authors reviewed and made substantial contributions to the manuscript.

## Information on Author Access to Data

Frank Lee and Cornelius Thiels had full access to all the data in the study and takes responsibility for the integrity of the data and the accuracy of the data analysis.

## Explanation of the Role of Funder(s)/Sponsor(s)

Funding organizations/sponsors had no role in design or conduct of this study nor decision to submit, review, or approval of this manuscript.

## Statement on Use of AI in Assisting with Writing/Editing Manuscript

No large language models (e.g. ChatGPT, Claude, Gemini, or Copilot) were used in the preparation of this manuscript.

## SUPPLEMENTAL CONTENT

**Figure A.1:**
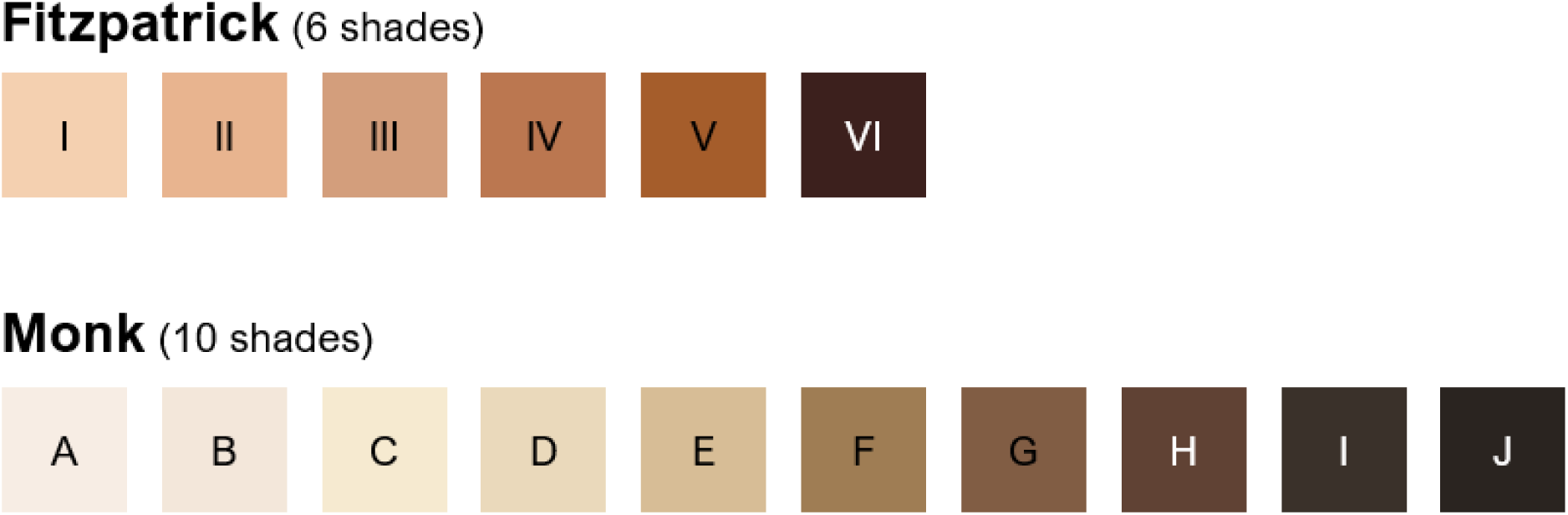
Reference guide for skin color labelling using Fitzpatrick and Monk scales

### Methods A.2: Utilized Vision Transformer Architecture

Vision Transformer (vit_large_patch16_224) is an architecture that leverages the power of the transformer model—originally popularized in natural language processing—to solve computer vision tasks.^40^ It splits an input image into 14×14 patches, treats each patch as a token, and uses multi-headed self-attention to capture relationships between these tokens at various positions. This approach allows the model to learn rich global context more efficiently compared to many conventional convolution-based networks. With its ability to process images as sequences, the Vision Transformer has demonstrated promising results across a variety of tasks, highlighting the versatility and potential of attention-based architectures.

**Table A.3:** Hyperparameter Search and Settings. A grid search approach was employed to optimize model hyperparameter using 10% of the training dataset. Learning rates were adaptively tuned via cosine annealing method ranging from 1×10⁻⁶ to 1×10⁻³ with a step size of 2×10⁻⁴. Batch sizes were selected in powers of two, ranging from 8 to 128. For each model, combinations of learning rate and batch size were systematically evaluated to identify those that maximized validation accuracy and minimized loss. The optimal set of hyperparameters was then used to train the final models. In the table below we provide the results for validation accuracy on a balanced validation set for different combinations’ of batch size and initial learning rate. We choose those values of hyperparameters which give us the highest validation accuracy for each model.

| Model | Batch Size | Learning rate | Validation Accuracy |
| --- | --- | --- | --- |
| Vision Transformer | 8 | $10^{-3}$ | 0.00 |
| | | $10^{-4}$ | 0.47 |
| | | $10^{-5}$ | 0.83 |
| | | $10^{-6}$ | 0.90 |
| | 16 | $10^{-3}$ | 0.32 |
| | | $10^{-4}$ | 0.49 |
| | | $10^{-5}$ | 0.84 |
| | | $10^{-6}$ | 0.90 |
| | 32 | $10^{-3}$ | 0.35 |
| | | $10^{-4}$ | 0.47 |
| | | $10^{-5}$ | 0.91 |
| | | $10^{-6}$ | 0.89 |
| | 64 | $10^{-3}$ | 0.12 |
| | | $10^{-4}$ | 0.52 |
| | | $10^{-5}$ | 0.90 |
| | | $10^{-6}$ | 0.86 |
| | 128 | $10^{-3}$ | 0.00 |
| | | $10^{-4}$ | 0.47 |
| | | $10^{-5}$ | 0.86 |
| | | $10^{-6}$ | 0.89 |

**Equations A.4**: Formulas for machine learning model performance

Accuracy (proportion of correctly classified images divided by total images):

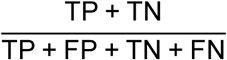

Precision a.k.a. Positive Predictive Value (proportion of true positive among all images classified as positive by the model):

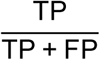

Recall a.k.a. Sensitivity:

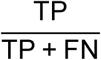

Specificity:

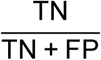

F1-score (measure of imbalance and is the harmonic mean of precision and recall):

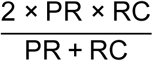

*\*TP true positive, TN true negative, FP false positive, FN false negative, PR precision, RC recall*

---

**Table A.5:** Surgical specialty case mix by geography.

| **Surgical Subspecialty** | **All Sites** | **RST** | **FLA** | **MCHS** | **ARZ** | **P** |
| --- | --- | --- | --- | --- | --- | --- |
| Orthopedics | 3,519 | 1,786<br>(22.3%) | 890<br>(37.4%) | 574<br>(30.7%) | 269<br>(18.6%) | <0.001 |
| Plastic surgery | 3,289 | 2,378<br>(29.7%) | 420<br>(17.7%) | 275<br>(14.7%) | 216<br>(15.0%) | <0.001 |
| General surgery | 2,298 | 1,400<br>(17.5%) | 340<br>(14.3%) | 351<br>(18.8%) | 207<br>(14.3%) | <0.001 |
| Neurosurgery | 1,370 | 749<br>(9.4%) | 152<br>(6.4%) | 256<br>(13.7%) | 213<br>(14.8%) | <0.001 |
| Otolaryngology | 891 | 526<br>(6.6%) | 210<br>(8.8%) | 41<br>(2.2%) | 114<br>(7.9%) | <0.001 |
| Gynecology | 709 | 390<br>(4.9%) | 87<br>(3.7%) | 165<br>(8.8%) | 67<br>(4.6%) | <0.001 |
| Urology | 592 | 371<br>(4.6%) | 62<br>(2.6%) | 17<br>(0.9%) | 142<br>(9.8%) | <0.001 |
| Vascular | 449 | 226<br>(2.8%) | 161<br>(6.8%) | 0 (0%) | 62<br>(4.3%) | <0.001 |
| Thoracic | 363 | 145<br>(1.8%) | 55<br>(2.3%) | 9 (0.5%) | 154<br>(10.7%) | <0.001 |
| Obstetrics | 222 | 38 (0.5%) | 0 (0%) | 184<br>(9.8%) | 0 (0%) | <0.001 |
| **Total** | **13,702** | **8,009** | **2,377** | **1,872** | **1,444** |  |

**Figure A.6:**
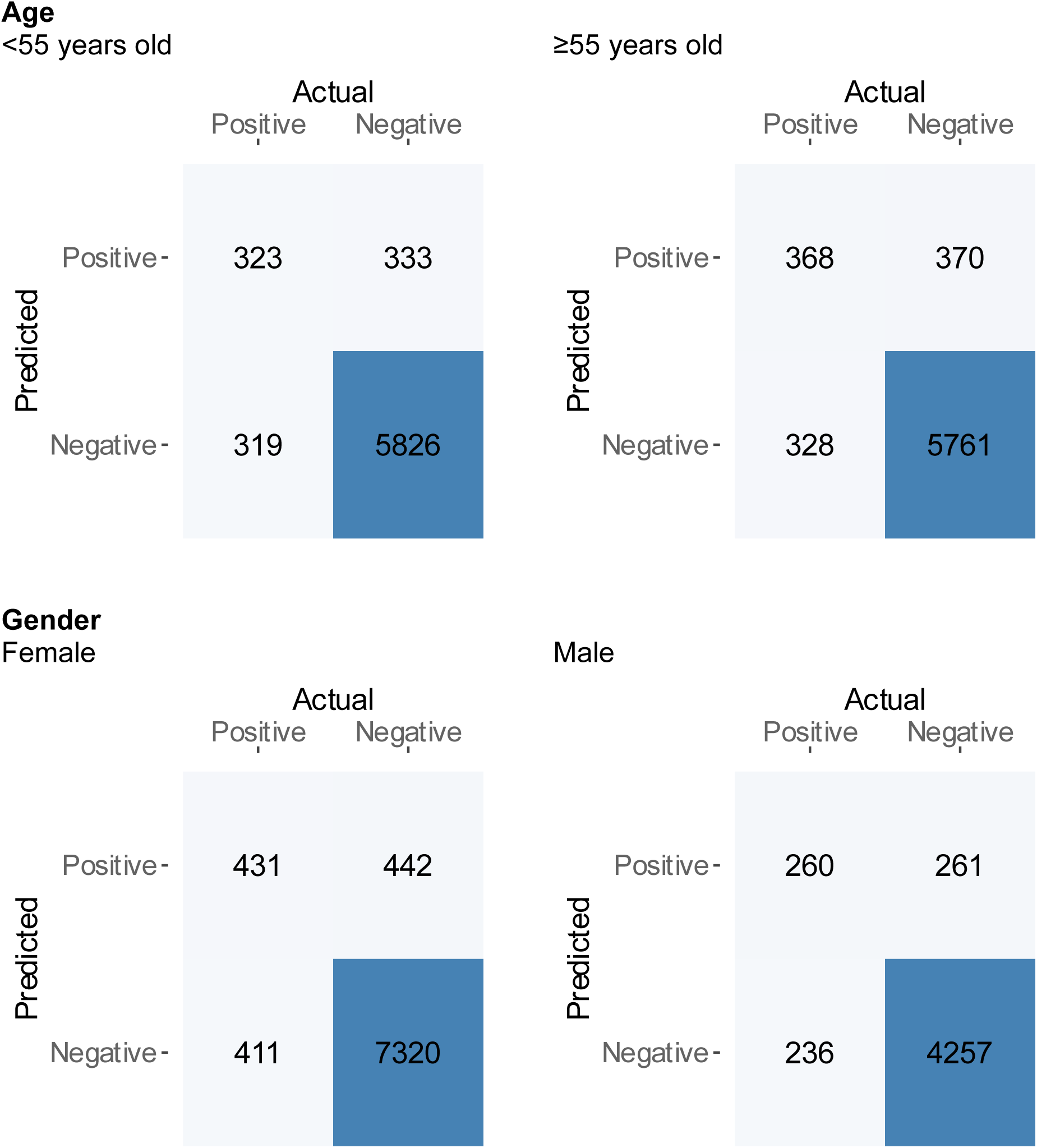
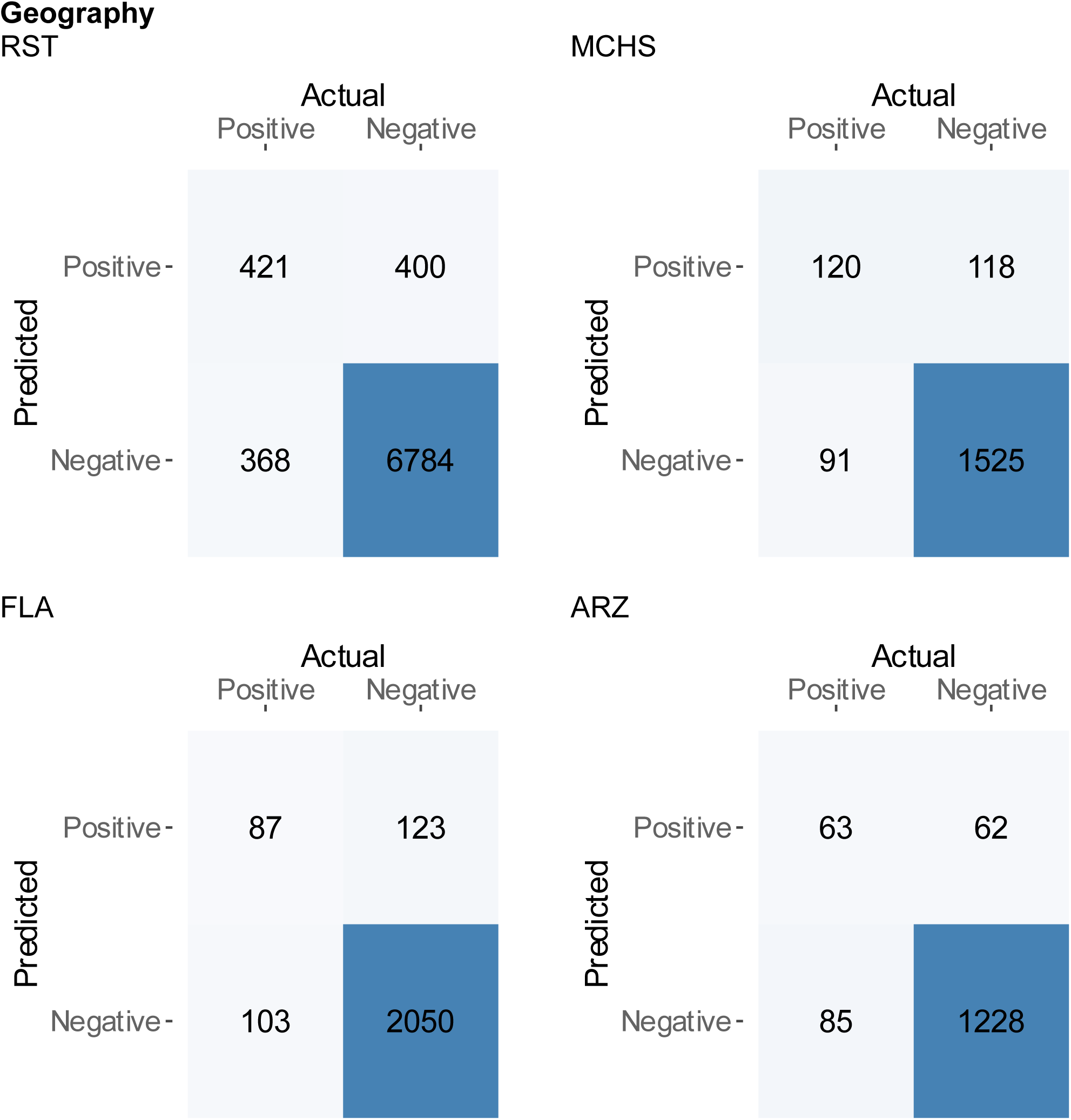
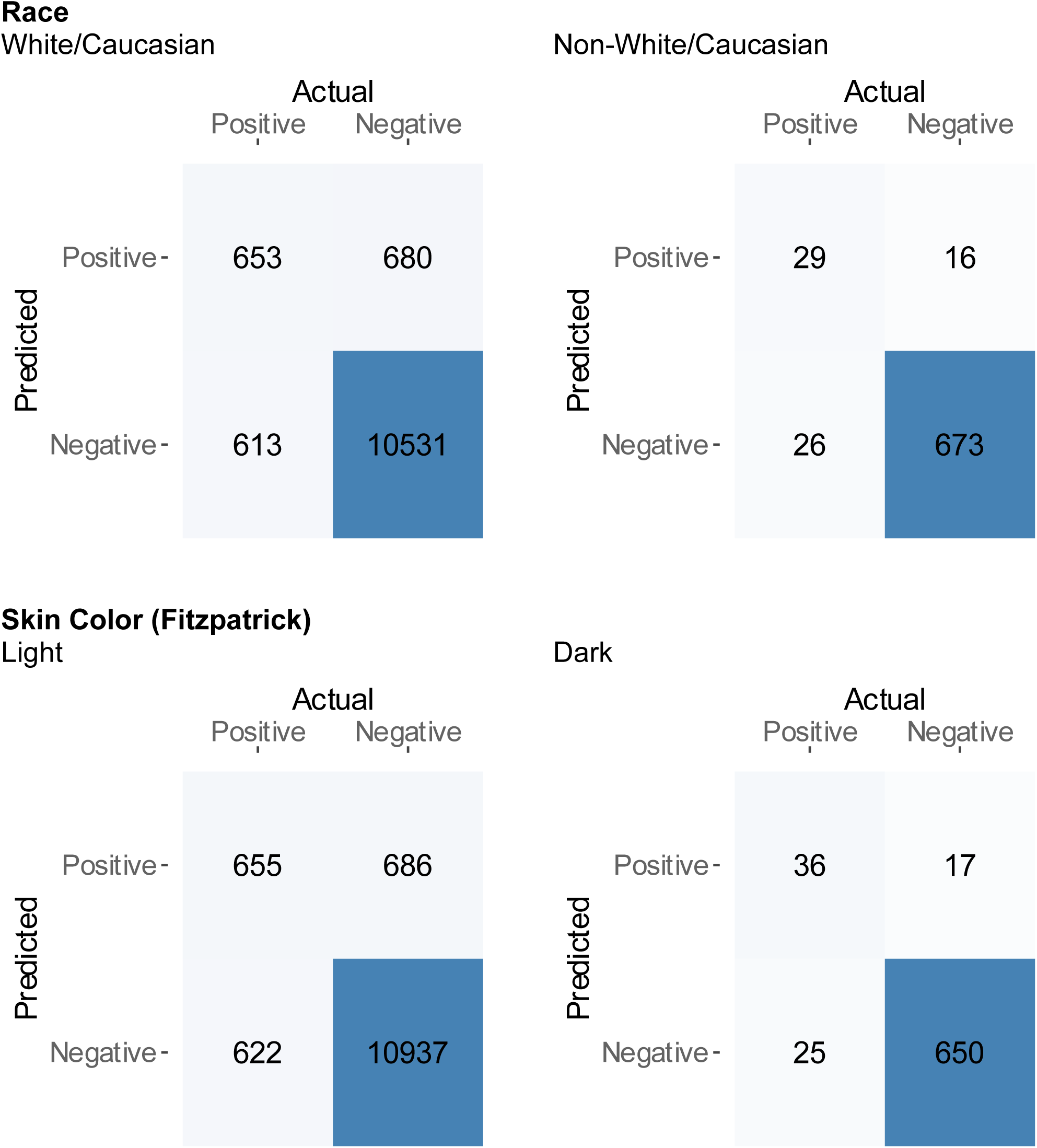
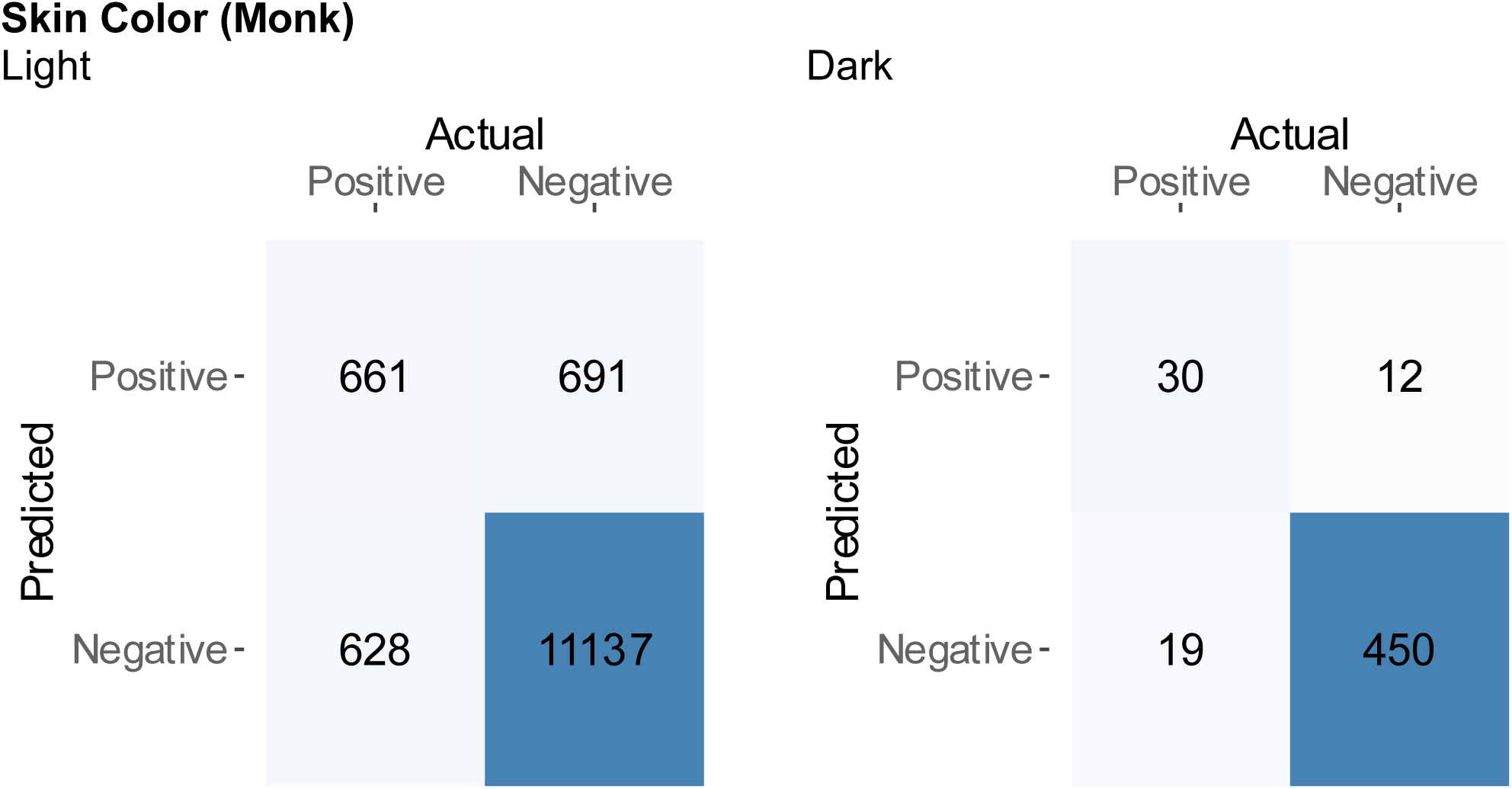
Confusion matrices analyzing subgroups during model development (threshold 0.416)

**Table A.7:**
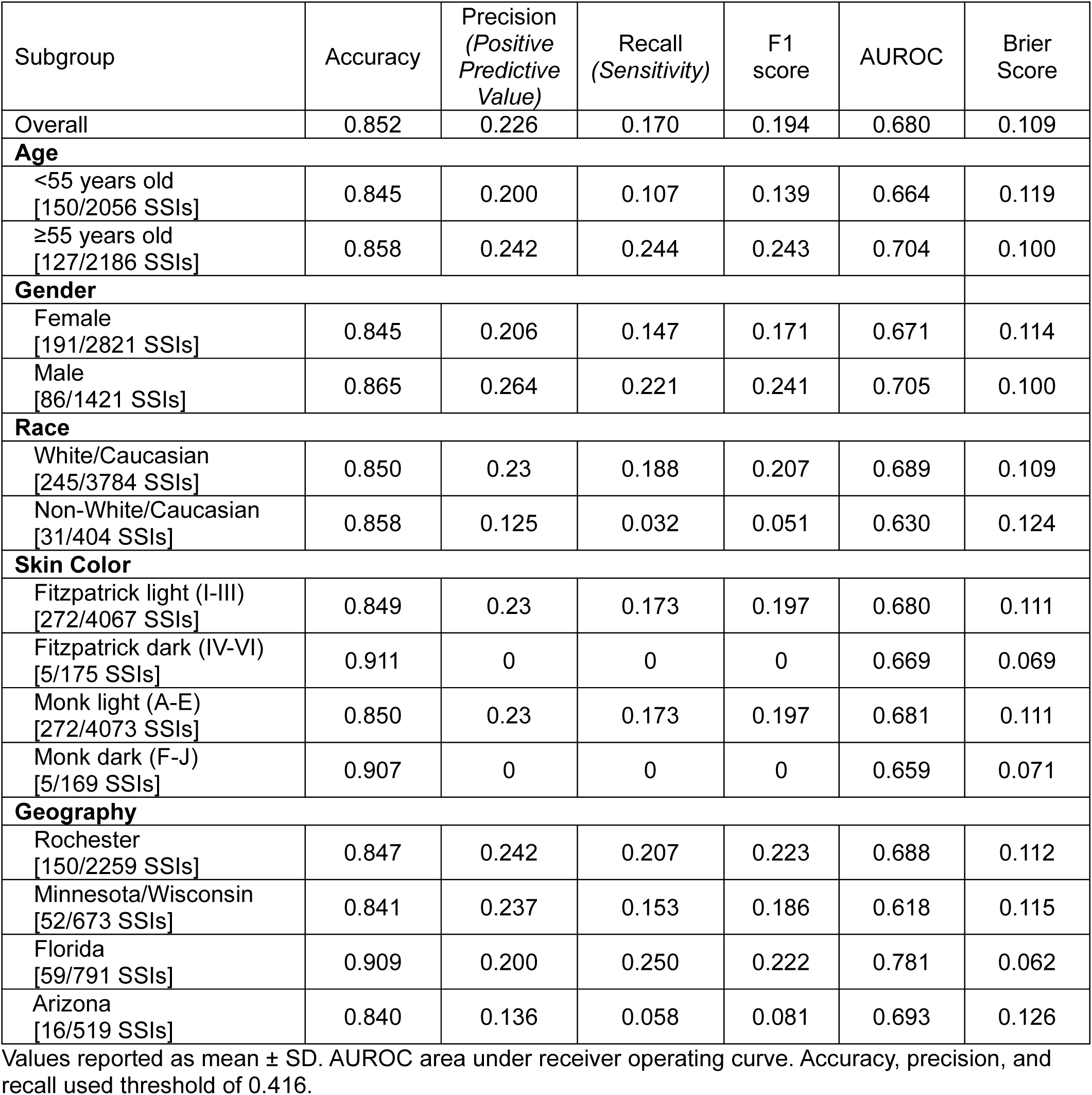
SSI detection performance metrics by subgroup during temporal validation.

| Subgroup | Accuracy | Precision<br>(Positive<br>Predictive<br>Value) | Recall<br>(Sensitivity) | F1<br>score | AUROC | Brier<br>Score |
| --- | --- | --- | --- | --- | --- | --- |
| Overall | 0.852 | 0.226 | 0.170 | 0.194 | 0.680 | 0.109 |
| **Age** |  |  |  |  |  |  |
| <55 years old<br>[150/2056 SSIs] | 0.845 | 0.200 | 0.107 | 0.139 | 0.664 | 0.119 |
| ≥55 years old<br>[127/2186 SSIs] | 0.858 | 0.242 | 0.244 | 0.243 | 0.704 | 0.100 |
| **Gender** |  |  |  |  |  |  |
| Female<br>[191/2821 SSIs] | 0.845 | 0.206 | 0.147 | 0.171 | 0.671 | 0.114 |
| Male<br>[86/1421 SSIs] | 0.865 | 0.264 | 0.221 | 0.241 | 0.705 | 0.100 |
| **Race** |  |  |  |  |  |  |
| White/Caucasian<br>[245/3784 SSIs] | 0.850 | 0.23 | 0.188 | 0.207 | 0.689 | 0.109 |
| Non-White/Caucasian<br>[31/404 SSIs] | 0.858 | 0.125 | 0.032 | 0.051 | 0.630 | 0.124 |
| **Skin Color** |  |  |  |  |  |  |
| Fitzpatrick light (I-III)<br>[272/4067 SSIs] | 0.849 | 0.23 | 0.173 | 0.197 | 0.680 | 0.111 |
| Fitzpatrick dark (IV-VI)<br>[5/175 SSIs] | 0.911 | 0 | 0 | 0 | 0.669 | 0.069 |
| Monk light (A-E)<br>[272/4073 SSIs] | 0.850 | 0.23 | 0.173 | 0.197 | 0.681 | 0.111 |
| Monk dark (F-J)<br>[5/169 SSIs] | 0.907 | 0 | 0 | 0 | 0.659 | 0.071 |
| **Geography** |  |  |  |  |  |  |
| Rochester<br>[150/2259 SSIs] | 0.847 | 0.242 | 0.207 | 0.223 | 0.688 | 0.112 |
| Minnesota/Wisconsin<br>[52/673 SSIs] | 0.841 | 0.237 | 0.153 | 0.186 | 0.618 | 0.115 |
| Florida<br>[59/791 SSIs] | 0.909 | 0.200 | 0.250 | 0.222 | 0.781 | 0.062 |
| Arizona<br>[16/519 SSIs] | 0.840 | 0.136 | 0.058 | 0.081 | 0.693 | 0.126 |

Values reported as mean ± SD. AUROC area under receiver operating curve. Accuracy, precision, and recall used threshold of 0.416.

